# Maternal Disrespect and Abuse by Health Care Providers among postpartum women attending Mizan-Tepi University Teaching Hospital, South West Ethiopia

**DOI:** 10.1101/2022.01.28.22269996

**Authors:** Dereje Sisay Kebede, Aklilu Mamo Dachew, Tafesse Lamaro Abota

## Abstract

**Background:** In Ethiopia, only 48% of all births occur at health facilities. Maternal disrespect and abuse by health care providers during child birth is one of the main reasons that affect health care seeking behaviors from health facilities. Thus, this study aims to assess magnitude of maternal disrespect and abuse during institutional delivery among postpartum women at Mizan -Tepi University Teaching Hospital (MTUTH), Bench Sheko Zone, South west Ethiopia

**Methods:** Institution-based cross-sectional study design was employed. A total of 409 postpartum women were enrolled using systematic random sampling technique between Augusts to September, in 2021. Pré-tested and structured questionnaire used for face-to-face interview. Data were coded, edited, and analyzed using SPSS version 25. Bivariate and multivariable logistic regression analyses were done. The presence and strength of association were determined using AOR with its 95% CI. Variables with P value of less than 0.05 were considered as statistically significant

**Results:** About half (52.1%) of the postpartum women experienced at least one category of maternal disrespect and abuse during institutional delivery. No formal education status during labor [AOR=2.216; 95% CI:1.266, 3.879], delivery during night shifts [(AOR=2.2; 95% CI: 1.306, 3.772], living in rural area [AOR=3.014;95% CI:1.630,5.572], number of health provider 2-3[AOR=4.101; 95%CI:2.70,6.23] and number of health provider above three[AOR=8.13; 95%CI:2.12,5.62] were found to be strongly associated with reporting maternal disrespect and abuse.

**Conclusions:** The study highlights that large proportion of mothers are experiencing maternal disrespect and abuse during institutional delivery in this study setting. Thus, appropriate interventions should be designed, focusing on supervision during night shifts, empowering childbearing women about their rights at health facilities and providing training for health care providers about respectful maternity care need to be emphatise.

## Background

Disrespectful care is a barrier to pregnant women who get public health care services. Disrespect and abuse in facility-based childbirth include physical abuse, non-dignified care, non-consented care, non-confidential care, discrimination, abandonment of care, and detention in facilities.^1^. One of the global agendas in Sustainable Development Goals (SDGs) is reducing maternal mortality to less than70 per 100,000 live births and ending preventable deaths of newborns and children under 5 years of age at least as low as 12 per 100 live births ^2^. An estimated 303,000 maternal and 2.7million neonatal mortality were occurred globally in 2015(3). Globally, one of the silent contributing factors for maternal morbidity and mortality is disrespect and abuse (D and A). For better maternal care service utilization, it is important to assess the existing prevalence of disrespect and abuse during child birth (4)(5). D & A during childbirth is a strong factor for choice of facility for the next childbirth(1)

Evenghough every region has advanced in maternal care service, the maternal mortality ratio is increasing in sub-Saharan Africa compared (6). By 2030, it is aimed to reduce the global maternal mortality ratio (MMR) to less than70 and focused on increasing antenatal care (ANC) coverage and facility-based childbirth as a key mechanism to reduce maternal mortality (3).

Ethiopia is one of the countries with the highest maternal mortality ratio in the world. According to the latest (2016) Ethiopian demographic and health survey report, maternal mortality ratio of the country was 353 per 100, 000 live births(5).

The magnitude of Disrespectful and abusive is more seen in many child birth facility settings particularly for underprivileged populations which violates their human rights and results in significant hindrance in accessing maternal care services(7). As a result, disrespect and abuse increases maternal and infant mortality by discouraging women from using facility-based childbirth (8).The burden of perinatal deaths is increasing now days particularly in developing country. So, developing the quality of care during childbirth, has been recognized as the most useful approach for decreasing perinatal death (9). According to Ethiopia Mini Demographic and Health Survey (EMDHS) 2019, the prevalence of institutional delivery was low (48%)(10).

Disrespect and abuse maternal care affects women’s rights to compassionate and respectful maternity care service utilization and impair their freedom which even leads to death as recognized by World Health Organization (11). In Ethiopia only 50% of births were attended by health professional (10). Deny of appropriate labor pain management, respectful care, fear of showing the body to health professionals, perceived cost of using a health facility during birth are all known as predisposing factor to low utilization of facility based birth (5)

In Ethiopia, only 48% of all births occur at health facilities. Even though few facilities based studies were conducted in Ethiopia (12) One of the main factor that affect health care seeking behaviors during child birth is maternal disrespect and abuse which resulted from health professional that seen in health facilities. Facilitating and ensuring compassionate and respectful maternity care services during labor and delivery is one of the most important interventions to maintain good maternal and newborn outcome.

However, compassionate and respectful maternity care has received much less attention both in practice and research. The reasons for failing to use skilled services during delivery have been studied a lot. But there is inadequate research on impact of disrespect and abuse of women during facility based deliveries that decreasing utilization of maternity services. Currently compassionate and respectful maternity care during labor and delivery considered as an important component of health care provider.. No study were conducted on prevalence of disrespect and abuse during child birth and its contributing factor in this setting. Therefore, the main aim of this study was to assess the magnitude of maternal disrespect and abuse and its associated factors among postpartum women who gave a birth at health facility.

## Methods and Materials

### Study design, setting, and participants

This study was conducted in MTUTH which is located in Bench-Sheko Zone of South West Ethiopia 574km from Addis Ababa, Ethiopia. MTUTH is the only teaching hospital in the zone which is populated by more than 800,000 people. It also give the service to adjacent zone including Kaffa, West Omo, Sheka and south part of Gambela region and refuges of South Sudan. The hospital has four patient wards: medical, surgical, pediatrics and obstetrics and gynecological wards. The obstetrics and gynecological ward consists of six waiting and fourteen postnatal beds with three delivery coaches, and two operation rooms’ tables. In addition, the ward has a total of 50 staffs (2 Obstetric and gynecologists, 8 general practitioners, 37 midwives, and 105 nurse). The average number of deliver per year for the hospital reaches around 2490.

### Source and study population

All mothers who delivered at MTUTU were source population. All postnatal mothers who were admitted to postnatal care (PNC) services during the study period considered study population

### Eligibility Criteria

All postnatal mothers who received delivery services for their last child birth at the study hospital and come to postnatal unit for their first postnatal services during the study period were included for this study. Postnatal mothers who were critically ill, unable to communicate at the time of data collection, refused to participate in the study, and they come to postnatal unit and those delivered by caesarian section were excluded.

### Sample size determination

The sample size required for this study was determined using a single population proportion formula considering the assumptions: Proportion of physical abuse (0.16), the woman’s right to;information, informed consent, and choice/ preferences was not protected(0.58), the woman’s confidentiality and privacy was not protected(0.45), non-dignified care(0.01), Stigma and discrimination(0.73), the woman was left without care/attention(0.39) and the woman was detained or confined against her will(0.02) during facility based child birth. These figures were taken from a previous study conducted in Five health centers of Addis Ababa(13) Level of confidence 95% and margin of error to be 5% (d = 0.05). The sample size for each form of disrespect and abuse was calculated as follows:

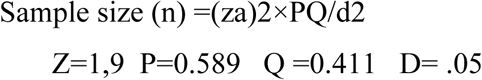

### Sampling procedure

In MTUTH the average number of deliveries per month in this hospital was 331 then during the two months of data collection period the number of deliveries was estimated to be a total of 662 deliveries. Systematic sampling technique was used for enrollment of 409 postnatal mothers who will be accessed for their first postnatal care services; by the assumption of: N (the estimated deliveries in two months period in the selected hospital which was s, and n (required minimum sample size = 409 which gives sampling fraction (k) of 2): k= N/n =>662 /409 =1.66 ≈ 2 To start data collection, the first two women who was admitted to PNC clinic for their first postnatal services were given numbers 1 or 2 and one of them from postnatal clinic was selected by lottery method to select the first participant (the first sampling unit). Then after every 2nd unit included until the required sample reached

### Study variables

#### Independent variables

Socio-demographic characteristics: Age, residence, income level, occupation, educational status

Obstetric characteristics: Parity, history of antenatal care follow-up, history of previous institutional delivery, place of birth, type of delivery

#### Dependent variable

Non respectful and abusive care: Physical abuse, non-consented care, non-confidential care, non-dignified care discrimination based on specific clients’ attributes, abandonment of care and detention in health facilitie

#### Data collection tool and procedures and quality

Data on participants’ demographic, individual related, obstetric history and provider related was collected using structured interviewer administrated questionnaire. For the sample participants, the purpose of the study and importance of participation was informed and verbal consent was ensured. Based on their willingness to participate in the study, a standardized structured interviewed questionnaire which is modified contextually was distributed to collect the data by trained four year regular nursing students.

The quality of data was assured by training of data collectors, further adjustment to the data collection tool was made after pre-testing with 5% of the sample size or 20 mothers at MTUTH which was not include in the study to improve clarity, understandability and simplicity of the message.

All of the questionnaires were checked for completeness and accuracy before, during and after the period of data collection. The collected data was again reviewed and checked for completeness before data entry. Data entry format template was prepared and programmed by principal investigator. Disrespect and abuse during childbirth was measured using 9 performance standards (categories of disrespect and abuse) and 35 verification criteria according to the new WHO framework of mistreatment of women during child birth (1)

#### Data processing, analysis, and interpretation

All the questionnaires were checked for completeness, coded and entered in to Epi Info version 7.1.2.0 and then transported to SPSS version 24 software package further analysis. Descriptive statistics such as mean, percentage and standard deviation were computed and presented, using frequencies and proportions for categorical variables, and summarized by mean and standard deviation for continuous variables. Frequencies of D&A components and subcomponents were also computed and reported in a table. Bi-variable logistic regression was done to determine the association between each independent variable and the outcome variables. Variables with a p-value of less 0.25 were considered candidates for the multivariable analysis to adjust the effect of confounders on the outcome variables. The degree of association between dependent and independent variables was determined using the odds ratio with confidence interval of 95% and p value of 0.05. The second analytical step used a stepwise backward model. The Homer and Lemeshow goodness-of-fit was applied to test for model fitness.

##### Ethics approval and consent to participate

This study was conducted in accordance with the Declaration of Helsinki. Ethical clearance obtained from Mizan-Tepi University research and community support officers. Before this survey, a formal letter was submitted to administrative office of Hospital. The study’s objective, benefit, and risks were explained to the participants before data collection and obtained written informed consent from all respondents. The study participants were assured of the attainment of confidentiality, and the information they give us will not be used for any purpose other than the study

## Results

### Background Characteristics of postpartum women attending Mizan-Tepi University Teaching Hospital, Benchi-Sheko Zone, South West Ethiopia

A total of 409 postpartum women were participated in this study with

’response rate of 100%. The average age of the participants was 31.3 years with a minimum and maximum age of 18 and 40 respectively. Majority of the respondents were 25 to 34 years old. About 44% of the study participants were from the Bench ethnic group. Approximately 45.7% of women were followers of Orthodox Christian religion (**Table 1**).

**Table 1:** The minimum sample size determination for each forms of D and A.

| Forms of D and A | Z | P | Q | D | N | Adding 10% for non-response Rate | Final |
| --- | --- | --- | --- | --- | --- | --- | --- |
| Physical abuse | 1.96 | 0.169 | 0.83 | 0.5 | 215 | 22 | 237 |
| Non consented care | 1.96 | 0.589 | 0.411 | 0.5 | 372 | 37 | 409 |
| Non confidentiality and privacy | 1.96 | 0.342 | 0.545 | 0.5 | 286 | 29 | 315 |
| Non dignified care | 1.96 | 0.013 | 0.987 | 0.5 | 197 | 20 | 217 |
| Stigma and discrimination | 1.96 | 0.733 | 0.26 | 0.5 | 293 | 29 | 322 |
| Neglected care | 1.96 | 0.39 | 0.61 | 0.5 | 365 | 37 | 402 |
| Detainment | 1.96 | 0.028 | 0.72 | 0.5 | 310 | 31 | 341 |

So, the large sample size 409 was taken in order to include all forms of disrespect and abuse.

**Table 1:** Socio demographic characteristics of mother in MTUTH, Bench Sheko Zone, South West Ethiopia, August 14-September 9, 2021 (n=409)

| Types of variable | Frequency | % |
| --- | --- | --- |
| Age |  |  |
| Less than 24 | 25 | 6.1 |
| 25-34 | 235 | 57.4 |
| ≥35 | 149 | 36.4 |
| Marital status |  |  |
| Married | 407 | 99.5 |
| Divorce | 2 | 0.5 |
| Residence |  |  |
| Urban | 185 | 45.2 |
| Rural | 224 | 54.8 |
| Ethnicity |  |  |
| Bench | 178 | 43.5 |
| Keffa | 129 | 31.5 |
| Others | 102 | 24.9 |
| Religion |  |  |
| Orthodox | 156 | 38.1 |
| Protestant | 187 | 45.7 |
| Others | 66 | 16.1 |
| Educational status |  |  |
| No formal education | 185 | 45 |
| Read and write | 188 | 45.9 |
| College and above | 37 | 9.1 |
| Occupation |  |  |
| House wife | 342 | 83.6 |
| Employed | 47 | 11.5 |
| Others | 20 | 4.9 |
| Family monthly income(ETB) |  |  |
| No income | 342 | 83.6 |
| <1000 | 14 | 3.4 |
| 1000-1500 | 12 | 2.9 |
| 1501-2000 | 36 | 8.8 |
| >2000 | 5 | 1.2 |

### Obstetric history of the postpartum women attending Mizan-Tepi University Teaching Hospital, Bench-Sheko Zone, South West Ethiopia, 2021

Out Of 409 respondents, 389(95%) had a history of ANC follow-up for their most recent delivery. Majority of the respondents had at least four visits for ANC service. The large proportion of women reported had previous history of institutional delivery for at least one child. Of the total sample, 279(68.2%) of mothers gave birth through spontaneous vaginal delivery. The vast majority, 336(82.2%) of women reported they had given birth preparedness education during ANC follow-up **(Table 2)**.

**Table 2:** Obstetric characteristics of mother in MTUTH, Bench Sheko Zone, South West Ethiopia August14-September 9, 2021 (n=409)

| Types of variable | Frequency | % |
| --- | --- | --- |
| Maternal ANC follow up |  |  |
| Yes | 389 | 95.1 |
| No | 20 | 4.9 |
| Number of ANC |  |  |
| <4 | 148 | 36.2 |
| ≥4 | 261 | 63.8 |
| gravidity |  |  |
| One | 21 | 5.1 |
| Two | 124 | 30.3 |
| Three | 182 | 44.5 |
| Four | 26 | 6.4 |
| Five and above | 56 | 13.7 |
| Total Number of delivery in health facility delivery |  |  |
| all | 221 | 54 |
| one | 24 | 5.9 |
| two | 38 | 9.3 |
| three | 109 | 26.7 |
| Four | 16 | 3.9 |
| Mode of recent delivery |  |  |
| SVD | 279 | 68.2 |
| SVD +Episiotomy | 127 | 31.1 |
| Vacuum delivery | 3 | 0.7 |
| Stay in health facility after delivery |  |  |
| 6-12 hours | 2 | 0.5 |
| 13-24 hours | 36 | 8.8 |
| >24 hours | 371 | 90.7 |
| Time of delivery |  |  |
| Night | 267 | 65.3 |
| Day | 142 | 34.7 |
| Complication during delivery |  |  |
| Maternal | 95 | 23.2 |
| Neonatal | 107 | 26.2 |
| Both | 9 | 2.2 |
| None | 198 | 48.4 |
| ANC follow up at the deliver hospital |  |  |
| yes | 87 | 21.3 |
| No | 322 | 78.7 |
| Mother's autonomy during labor and delivery |  |  |
| Yes | 213 | 52.1 |
| No | 196 | 47.9 |
| Sex of main provider conducting delivery |  |  |
| male | 190 | 46.5 |
| female | 219 | 53.5 |
| Number of provider who assisted during delivery |  |  |
| One | 42 | 10.3 |
| Two | 233 | 57 |
| Three or four | 134 | 32.8 |
| Information on birth preparedness given during ANC follow up |  |  |
| yes | 336 | 82.2 |
| No | 73 | 17.8 |

### Magnitude of maternal disrespect and abuse among postpartum during institutional delivery at MTUTH in Bench-Sheko Zone, South West Ethiopia

Of the total sample, half (52.1%) of the postpartum women reported that they have experienced at least one form of Disrespect and Abuse during facility based Childbirth. Based on verification criteria for categories of D and A, we counted mothers who faced at least one condition among the possibilities. Accordingly, the most commonly experienced form of D and A was ineffective communication between maternity care providers and women during labor and delivery213 (52%). **(Table 3)**

**Table 3:**
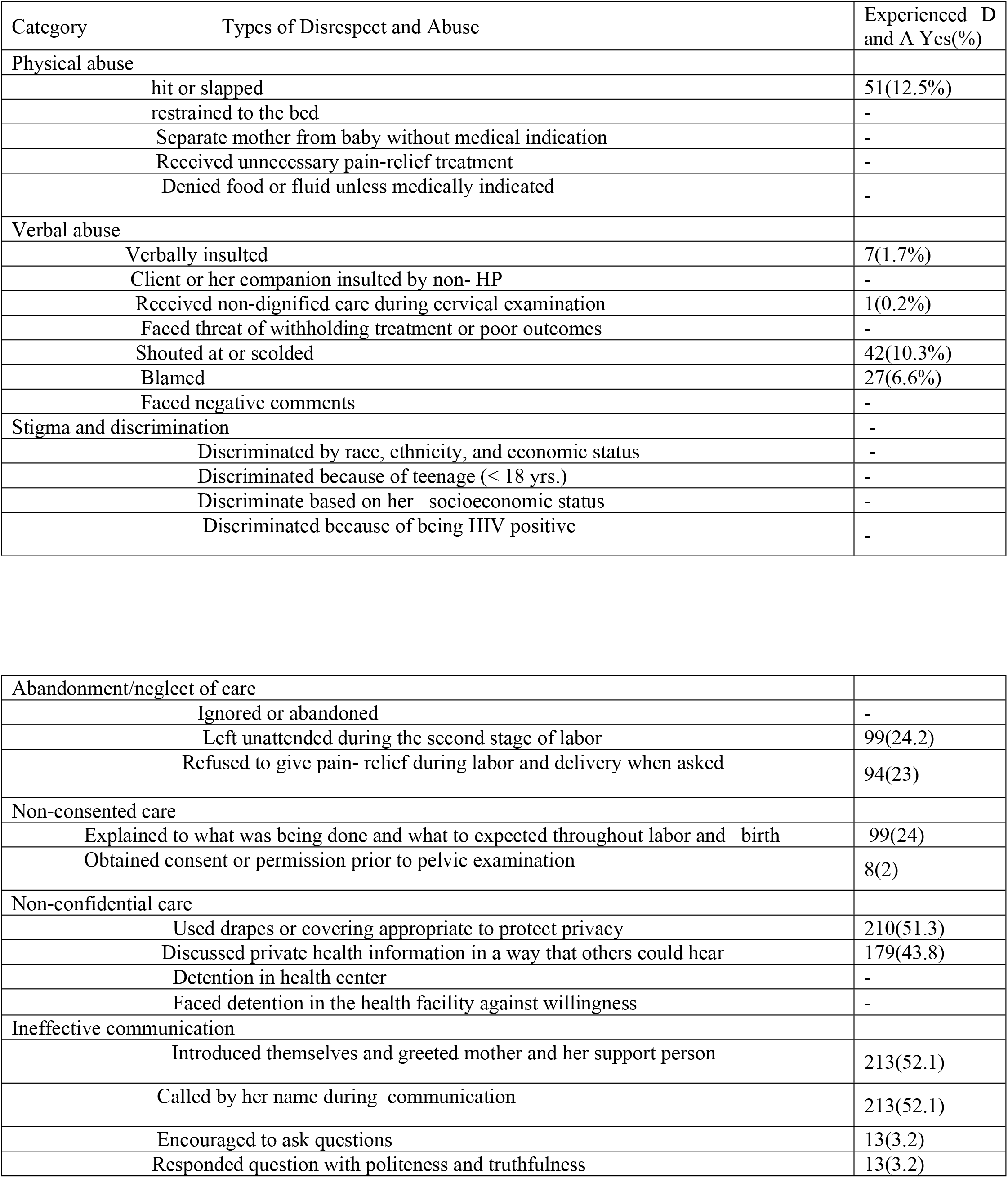
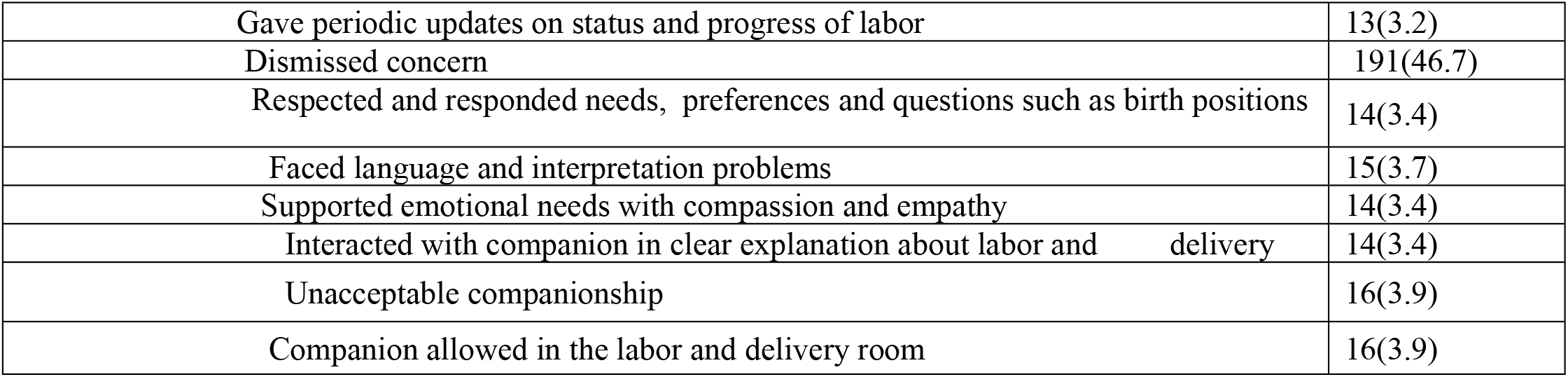
Prevalence of disrespect and abuse during childbirth by categories, MTUTH, Bench Sheko zone, South West Ethiopia, Augest 14-September 9, 2021G.C

| Category | Types of Disrespect and Abuse | Experienced D and A Yes(%) |
| --- | --- | --- |
| Physical abuse |  |  |
|  | hit or slapped | 51(12.5%) |
|  | restrained to the bed | - |
|  | Separate mother from baby without medical indication | - |
|  | Received unnecessary pain-relief treatment | - |
|  | Denied food or fluid unless medically indicated | - |
| Verbal abuse |  |  |
|  | Verbally insulted | 7(1.7%) |
|  | Client or her companion insulted by non- HP | - |
|  | Received non-dignified care during cervical examination | 1(0.2%) |
|  | Faced threat of withholding treatment or poor outcomes | - |
|  | Shouted at or scolded | 42(10.3%) |
|  | Blamed | 27(6.6%) |
|  | Faced negative comments | - |
| Stigma and discrimination |  | - |
|  | Discriminated by race, ethnicity, and economic status | - |
|  | Discriminated because of teenage (< 18 yrs.) | - |
|  | Discriminate based on her socioeconomic status | - |
|  | Discriminated because of being HIV positive | - |
| Abandonment/neglect of care |  |  |
|  | Ignored or abandoned | - |
|  | Left unattended during the second stage of labor | 99(24.2) |
|  | Refused to give pain- relief during labor and delivery when asked | 94(23) |
| Non-consented care |  |  |
|  | Explained to what was being done and what to expected throughout labor and birth | 99(24) |
|  | Obtained consent or permission prior to pelvic examination | 8(2) |
| Non-confidential care |  |  |
|  | Used drapes or covering appropriate to protect privacy | 210(51.3) |
|  | Discussed private health information in a way that others could hear | 179(43.8) |
|  | Detention in health center | - |
|  | Faced detention in the health facility against willingness | - |
| Ineffective communication |  |  |
|  | Introduced themselves and greeted mother and her support person | 213(52.1) |
|  | Called by her name during communication | 213(52.1) |
|  | Encouraged to ask questions | 13(3.2) |
|  | Responded question with politeness and truthfulness | 13(3.2) |

|  |  |
| --- | --- |
| Gave periodic updates on status and progress of labor | 13(3.2) |
| Dismissed concern | 191(46.7) |
| Respected and responded needs, preferences and questions such as birth positions | 14(3.4) |
| Faced language and interpretation problems | 15(3.7) |
| Supported emotional needs with compassion and empathy | 14(3.4) |
| Interacted with companion in clear explanation about labor and delivery | 14(3.4) |
| Unacceptable companionship | 16(3.9) |
| Companion allowed in the labor and delivery room | 16(3.9) |

### Factors associated with experiencing maternal Disrespect and Abuse among postpartum women attending Mizan-Tepi University teaching hospital, Benchi-Sheko Zone, South West Ethiopia

In bivariate analysis, Religion, Occupation, Monthly income, Number of delivery in health facility, Stay in hospital after delivery, Number of health provider attending the delivery, Time of delivery, Sex of main provider conducting the delivery and Involvement in birth plan with provider were found to be significantly associated with maternal disrespect and abuse. After controlling for other factors, religion, educational status and gender of the heath care provider attending delivery were significantly associated with disrespect and abuse. The odds of maternal disrespect and abuse among mothers who gave birth at night shifts were 2.2 times higher than those who delivered at day shifts [AOR=2.21; 95% CI:1.306, 3.772]. The likelihood of reporting disrespect and abuse among mothers who have no formal educational status during labor and delivery were 2.20 times higher than those who can read and write [AOR=2.216; 95% CI:1.266, 3.879]. Similarly, the odds ratio of disrespect and abuse among respondent who lives in rural area were 3.014 times higher than those who were live in urban [AOR=3.014, 95% CI:1.630,5.572] (Table 4).

**Table 4:**
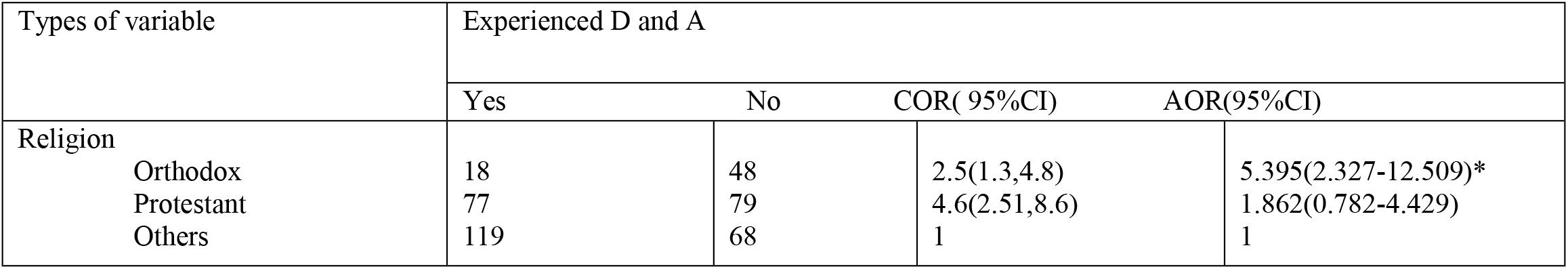
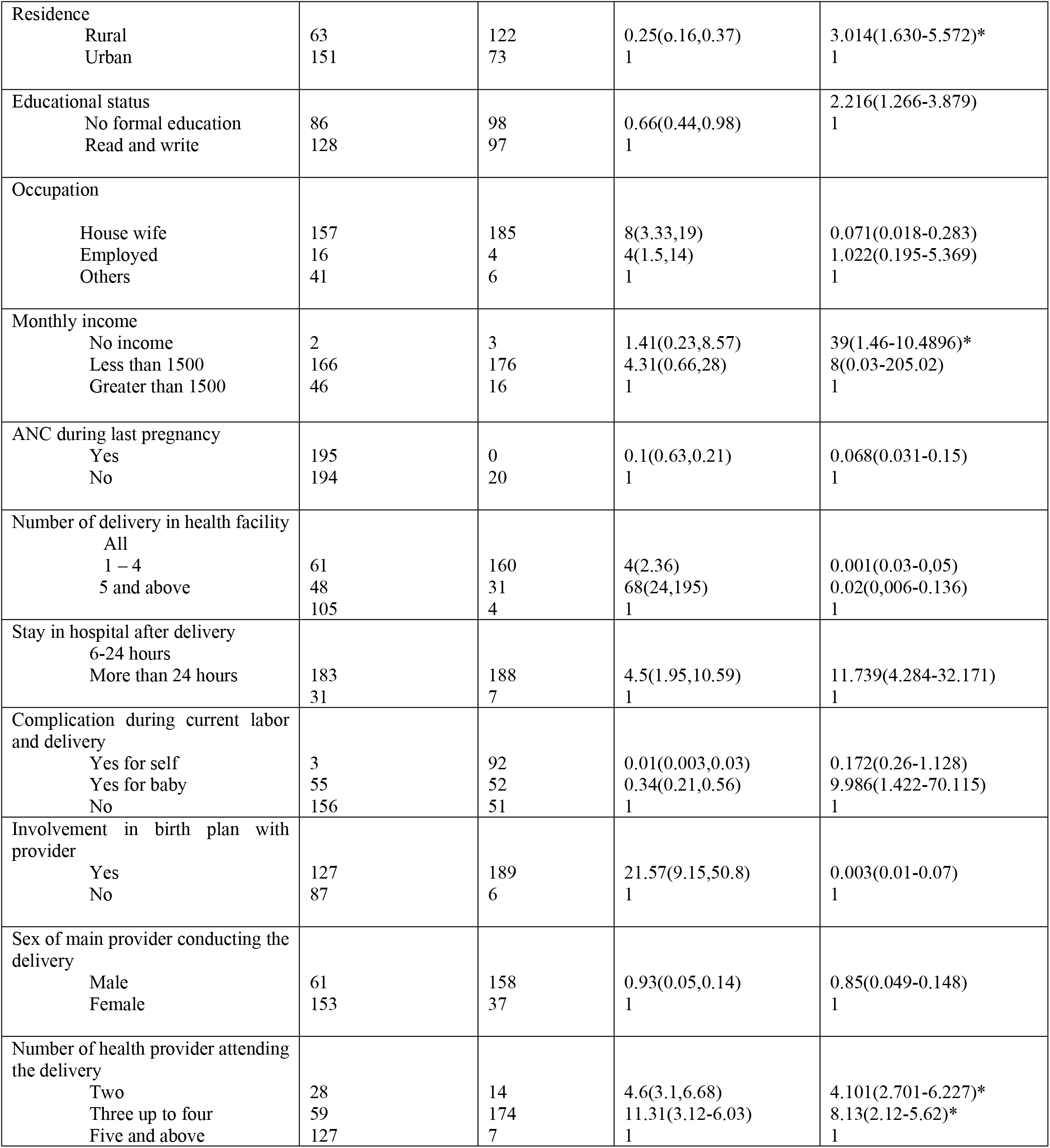
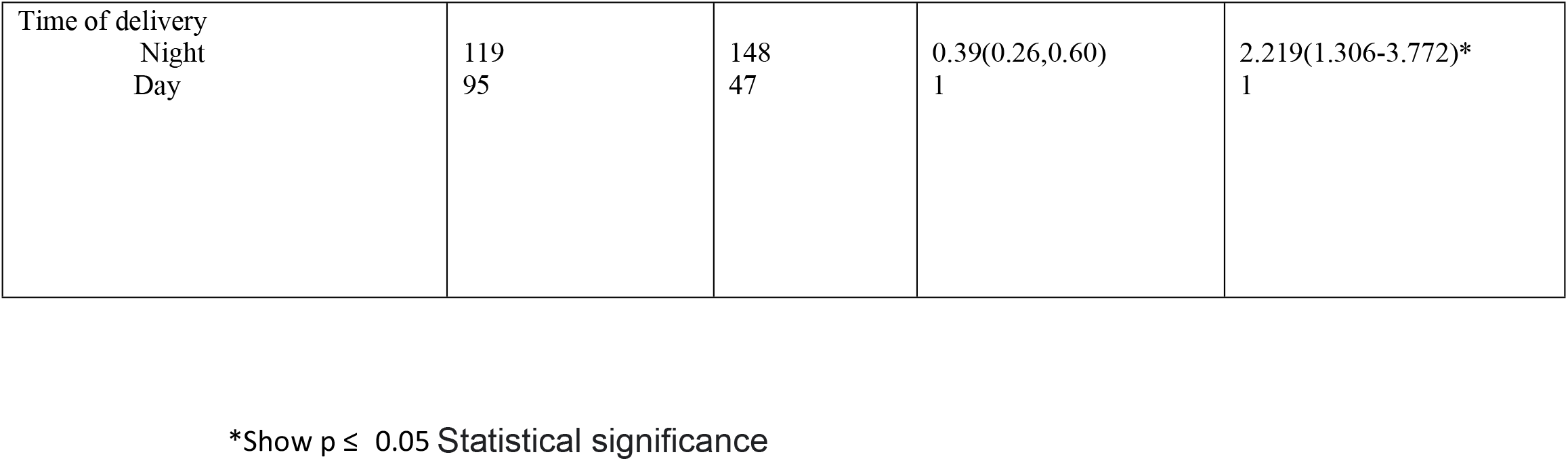
Bi-variable and Multi variable logistic regression analysis of disrespect and abuse and its explanatory variables (n=409)

## Discussion

This study investigated magnitude of maternal disrespect and abuse among postpartum women during facility based child birth in MTUTH. In the present study, overall magnitude of disrespect and abuse during labor and delivery was found to be high. The manifestation of disrespect and abuse measured using different entities: Ineffective communication, non-consented care, non-confidential care, verbal abuse, abandonment/neglected care and physical abuse. The current finding revealed that magnitude of disrespect and abuse is lower than findings from a study conducted in Arba Minch town (98.9%) in Ethiopia and Nigeria (98%), Bahir Dar (67.1%) and Addis Ababa (75.3%) (5,14),

This discrepancy might be due to the difference of study settings. The previous studies include a hospital and health centers in Ethiopia and Nigerian study at teaching hospital. The current study also use nine categories of disrespect and abuse to measure its extent. In line with this, ineffective communication and unacceptable companionship were included which are crucial to respectful maternity care during labor and delivery. The previous studies used the same definitions for the categories of disrespect and abuse as the present study, but used fewer items: whereas 35 items were used in the present study, the studies in Bahir Dar and Addis Ababa used 25, 23 items respectively (5,14).

According to this study, ineffective communication is the most commonly experienced component of disrespect and abuse and its prevalence was 52%. This showed that above half of the women faced poor communication that reflects women’s social and cultural needs, where relevant to labor and childbirth, despite communication being referred to as a core component of respectful maternity care. This might be due to maternity care providers give less attention to effective communication than other categories of D and A. The most commonly experienced form of ineffective communication was provider did not introduce himself/herself to mother and her companion and greet in respectful manner. This might be due to health care providers that consider respectful greeting is not as such important.

The second commonly reported types of D and A was non confidential care and verbal abuse (51%). This figure was lower than study conducted in Malawi (15). This discrepancy might be due to data collection method. The previous study was from direct observations of labor and delivery. Asking women for agreement is an important measure of showing respect for the laboring mother. In this study, 24% of laboring mother received non-consented care. This might be due to women didn’t know they had rights to be asked their consent before any procedures. However, this finding was lower than the same study conducted in Tanzania (16). This inconsistency might be due to difference in health police and implementation program. The statement of the universal rights of childbearing women states that healthcare providers must protect the patient’s privacy and confidentiality during any procedure and when handling a woman’s information (11). In contrast, this study revealed that 51% of women had been provided care in a non-confidential manner. This could be due to the lack of appropriate physical barriers like curtains at health facilities and/or poor understanding of the importance of confidentiality during childbirth among healthcare providers. This finding is high from the study which was conducted in urban Tanzania and Kenya(17,18).This difference might be due to data collection methods. The previous studies found data from direct observation of mothers during labor and delivery.

According to these findings, the other category of disrespect and abuse experienced by women was verbal abuse which is similar with non-confidential care (51%). This discrepancy might be due to fact that there is socio cultural and socio economical difference that affect mothers’ behavior and their reactions during labor and delivery. But, this finding was higher than the study conducted in Peru (19). This difference might be due to data collection method and study setting.

Similarly the other category of disrespect and abuse reported in this study was abandonment/neglected care during labor which accounts for24.2 %. This figure is lower than the study conducted in Kenya to measure disrespect and abuse during facility based child birth using direct observation(20).The difference may be due to hawthorn effect, in which behavior under study changes because the actors know they are being observed. This could be due to lack of empathy by health care providers for continuous caring laboring mothers.

The other category of disrespect and abuse reported in this study was physical abuse which accounts the prevalence of 12.5 %. This figure is lower than the studies conducted in Nigeria, Addis Ababa and Bahir Dar, Ethiopia (5,14,21). The discrepancy might be due to good commitment of health care professionals against physical abuse. This finding is similar with studies conducted in Pakistan and India on women’s experiences of mistreatment during facility based childbirth (22,23)

The odds of maternal disrespect and abuse among mothers who gave birth at night shifts were 2.2 times higher than those who delivered at day shifts [AOR=2.21; 95% CI:1.306, 3.772].This finding in line with study conducted in Kenya (17). In line with this, qualitative study conducted in Ethiopia revealed disrespectful and abusive care were more common during night shifts(4). The possible justification might be due to workload and dissatisfaction by health care providers.

The likelihood of reporting disrespect and abuse among mothers who have no formal educational status during labor and delivery were 2.20 times higher than those who can read and write [AOR=2.216; 95% CI:1.266, 3.879. This finding is higher than the studies conducted in Addis Ababa and Bahir Dar, Ethiopia (13,21)The discrepancy might be due to women with no formal educational may not know the statement of the universal rights of childbearing women that states healthcare providers must protect the patient’s privacy and confidentiality during any procedure and the right of handling a woman’s information confidentially

The odds ratio of disrespect and abuse among respondent who lives in rural area were 3.014 times higher than those who were live in urban [AOR=3.014, 95% CI:1.630,5.572] This figure is higher than the studies conducted in Nigeria, Addis Ababa and Bahir Dar, Ethiopia (5,14,21).The discrepancy might be due to women who lives in rural area may not get access to health police and implementation program that support and stand for right of women who delivery at facility based child birth as well as do not Know the statement of the universal rights of childbearing women that states healthcare providers must protect the patient’s privacy and confidentiality during any procedure and the right of handling a woman’s information confidentially

Low maternal family income status was also a factor for reporting disrespectful care and abuse. The finding is consistent with study set in Addis Ababa which indicates poor women were more disrespected and abused than the rich (14). evidence from Kenya reveals the rich women received care earlier compared to the poor, despite the seriousness of the medical condition(20).

The gender of health care providers was associated with experiencing maternal disrespect and abuse. The women who gave birth and assisted by female health care provider were more likely to have had disrespectful care and abuse during labor and delivery.

In the current study, maternal religion was associated with reporting of disrespectful care and abuse. A mother whose religion was orthodox were more likely disrespected and abused than those mothers whose religion was protestant. This result agrees with the study conducted in Tanzania (16). The interpretation of this finding remains very complex and needs further interpretations. In the present study, duration of Stay in hospital after delivery attended by health care providers was found to be indictors of experiencing with disrespectful care and abuse[AOR= 11.739 95% .CI (4.284-32.171]. This study agree with that of study conducted in Kenya (17).

### The Strengths and Limitations

The study tried to measure nine categories of D and A based on the new WHO framework using 35 verification criteria. So, it reduces underestimate of disrespectful and abusive practices during childbirth. The study also collected data from first postnatal mother visitors. This it may tackle information and recall biases. Since study relied on self-report, it does not provide an objective measure of the frequency of poor and abusive care in facility. The study also not supported by qualitative study to get information about D and A from community leader prospective and maternity care user prospective

### Conclusions and recommandations

In this study, women receiving labor and delivery care at Mizan Tepi University Teaching Hospital are experiencing maternal disrespect and abuse to a higher extent suggesting intervention. Ineffective communication was found as one of the commonest types of disrespectful care at this Teaching Hospital. Abusive and disrespectful care at this hospital is critical concern, which urges attention to promote women-friendly care for all women. This maternal disrespect and abuse could result in low use of health care service which needs immediate measures by health care managers. This study in general indicates the need for a more integrated intervention including empowering all women of childbearing age about their rights at this hospital, the economic empowerment of women, the type of care they deserved and providing training for all health care providers both on job and during their basic trainings Should be prioritized in order to improve access to medical services and combat disrespect and abusive care The problame is particularly high among uneducated women and women who are from rural areas. Women’s right to information and informed consent was the most frequently violated while Stigma and discrimination in hôpital facilites was reported by none of the respondents. Some of the misconducts might be related dis satisfaction of staffs and also needs serious attention by the Heath managers of the institutions.

Further research to disclose barriers and facilitators of respectful and dignified maternal care in accordance to providers and the health system is needed to design and implement effective interventions to maintain respectful maternity care in area of limited resources

## Data Availability

xxx

## Acknowledgments

We are grateful to Mizan-Tepi university teaching hospital administration and staffs for their marvelous collaboration, College of Medicine and Health Sciences, for ethical clearance. Finally, we would also like to thank data collectors, supervisors, study participants, and staff for their honest and kind cooperation and all women who shared their stories and time for this research.

## Authors’ contributions

Conceptualization: Dereje Sisay Kebede,

Data curation: Dereje Sisay Kebede, Aklilu Mamo Dachew, Tafesse Lamaro Abota

Formal analysis: Dereje Sisay Kebede,

Funding acquisition: Dereje Sisay Kebede,

Investigation: Dereje Sisay Kebede,

Methodology: Dereje Sisay Kebede, Aklilu Mamo Dachew, Tafesse Lamaro Abota

Project administration: Dereje Sisay Kebede

Resources: Dereje Sisay Kebede

Software: Dereje Sisay Kebede, Aklilu Mamo Dachew, Tafesse Lamaro Abota

Supervision: Dereje Sisay Kebede

Writing – original draft: Dereje Sisay Kebede

Writing – review & editing: Dereje Sisay Kebede, Aklilu Mamo Dachew, Tafesse Lamaro Abota

